# Towards Electronic Health Records-Based Paediatric Growth References: Results from the SwissPedGrowth Project

**DOI:** 10.64898/2026.08.28.26361619

**Authors:** Lorenz M. Leuenberger, Yara Shoman, Franco Romero, Mari Sasaki, Xeni Deligianni, Nicole Goebel, Rebeca Mozun, Julia A. Bielicki, Marie-Anne Burckhardt, Christoph Saner, Valérie Schwitzgebel, Michael Hauschild, Franziska Righini Grunder, Pascal Müller, Luregn J. Schlapbach, Oskar G. Jenni, Ben D. Spycher, Claudia E. Kuehni, Fabiën N. Belle, SwissPedHealth consortium

## Abstract

**BACKGROUND:** We used anthropometric data from electronic health records (EHRs) of Swiss children’s hospitals to evaluate growth references and estimate centile curves.

**METHODS:** We received EHRs extracted from seven Swiss children’s hospitals and analysed two samples: all children with a height, weight, body mass index (BMI), or head circumference recording, and a subsample restricted to children without diseases potentially affecting growth, weighted to represent the general population. We calculated mean z-scores based on the World Health Organization growth references adopted for Switzerland in 2011 (CH-WHO 2011) and current Swiss growth references (Swiss 2026). We estimated sex-specific centile curves in the subsample using generalised additive models for location, scale, and shape.

**RESULTS:** We included 213,868 children with height, 448,002 with weight, 209,244 with BMI, and 67,397 with head circumference recordings. Mean z-scores in the ‘all children’ sample were (CH- WHO 2011; Swiss 2026): height (0.10; -0.19), weight (0.16; -0.09), BMI (0.04; -0.07), head circumference (-0.28, -0.28); and in the subsample: height (0.34; 0.00), weight (0.27; 0.01), BMI (0.18; 0.05), and head circumference (0.04; 0.01). The 50^th^ height, weight, BMI, and head circumference centiles of girls and boys in the subsample closely followed those of Swiss 2026, with slightly wider 3^rd^ and 97^th^ centiles in infancy and adolescence.

**CONCLUSION:** Height, weight, BMI, and head circumference centiles aligned well with the Swiss 2026 growth references in Switzerland, demonstrating that hospital EHRs could contribute to future growth references.

## INTRODUCTION

Growth references and their corresponding percentile curves of body length/height, weight, body mass index (BMI), and head circumference are an important tool for paediatricians, enabling growth monitoring, identifying abnormal growth patterns and underlying health conditions. The utility of growth references for clinics and public health depends on the representativity of the paediatric population; poorly fitting references may lead to misclassification of stature or weight [1]. To represent local populations, many European countries have constructed national growth references [2–5].

In Switzerland, growth references representative of the local population of Central and Eastern Switzerland have been published in 2019 by the Paediatric Endocrinology Centre Zurich (PEZZ), a private practice of paediatric endocrinologists [6]. These references fitted to school children from the canton of Zurich better than previous World Health Organization (WHO) references used in Switzerland since 2011 (CH-WHO 2011) [1, 7]. After expanding data collection to be more representative of the entire country, PEZZ produced new growth references in 2025, which were recommended by pädiatrie schweiz (Swiss Society of Paediatrics) for clinical use in 2026 (Swiss 2026) [8, 9].

Traditional data collection for growth references is resource intensive. The use of electronic health records (EHRs) may present an attractive alternative. In 2019, new growth references in France were constructed from EHRs extracted from primary care practices [2], demonstrating the potential of routinely collected data for this purpose. In Switzerland, the Swiss Personalized Health Network (SPHN) built an interoperable framework for multi-institutional sharing of EHRs across Swiss hospitals, including a paediatric data stream (SwissPedHealth) with over 750,000 children [10].

SwissPedHealth allowed us to compare anthropometric data from routine care with previous (CH- WHO 2011) and contemporary (Swiss 2026) Swiss growth references, and to assess the potential of hospital EHRs for conducting growth research and constructing future growth references by evaluating how well anthropometric data of children visiting Swiss paediatric hospitals fitted the Swiss growth references. We also modelled growth references based on hospital data.

## MATERIALS AND METHODS

### Study design

The SwissPedHealth paediatric data stream includes seven paediatric hospitals in Switzerland [10]: Basel, Bern, Geneva, Lausanne, Lucerne, St. Gallen, and Zurich. Participating hospitals extracted EHRs of inpatient and outpatient encounters from 2017–2023 for children aged <20 years at the time of admission. SwissPedGrowth is nested in SwissPedHealth and was approved by the cantonal ethics committee Bern (KEK Bern 2023-00022): It permitted re-use of healthcare data from patients who provided general hospital consent (01.01.2017–31.12.2025), as well as from those who were informed of the general consent and did not actively refuse (01.01.2017–28.02.2023). For the present analysis, we included data from all children with available information on age and sex and at least one height, weight, BMI, or head circumference recording obtained before age 18 years.

### Data extraction and preparation

The data extraction and preparation steps of SwissPedGrowth have been described previously [11]. A detailed description can be found in the supplementary material. Hospitals extracted demographic, administrative, and clinical data and linked the Neighbourhood Index of Socioeconomic Position (Swiss-SEP), an area based index ranging from 0 (low SEP) to 100 (high SEP), to the patient’s home address using a standard operating procedure [13].We cleaned the anthropometric data using a self- developed algorithm and the pre-existing growthcleanr package in R [11, 16], identifying and correcting unit and decimal point errors, and excluding duplicates and biologically implausible values (height and head circumference z-scores <-5 or >5; weight and BMI z-scores <-5 or >8). To create a dataset with BMI values, we matched the closest height recording to each weight recording, allowing for a maximum interval of 30 days, as previously described [11, 15]. We categorised International Classification of Disease 10^th^ version (ICD-10) diagnoses according to their potential influence on anthropometric parameters, based on exclusion criteria from previous growth studies and the expert opinion of a panel of paediatric growth specialists (Supplementary Table S1) [5, 17]. We grouped nationality as Swiss, Northern/Western European, Southern/Eastern European, and Other (non- European).

### Samples of children for analysis

We defined two analytic samples: an ‘all children’ sample, and a ‘normal growth and weighted’ subsample. The ‘all children’ sample comprised all children with at least one height, weight, BMI, or head circumference recording. For the ‘normal growth and weighted’ subsample, we excluded anthropometric measurements that were affected by disease, see above and supplementary material, as well as children without information on nationality or Swiss-SEP. We then weighted each subsample (height, weight, BMI, and head circumference) to represent the age, sex, nationality, and Swiss-SEP distribution of the general paediatric population aged <18 years using iterative proportional fitting (raking) with the *survey* package in R, based on census data from the Swiss Federal Statistical Office [18]. We evaluated the weighting by calculating standardised differences (Cohen’s h [19]) between the SwissPedGrowth subsample and the general paediatric population before and after weighting, interpreting standardised differences of <0.1 as small [19].

### Statistical analysis

We calculated z-scores for height, weight, BMI, and head circumference using the age-specific lambda-mu-sigma (LMS) values from the CH-WHO 2011 and Swiss 2026 growth references. For BMI analyses, the two growth references overlapped in children aged 0–2 years, because the Swiss 2026 BMI reference re-used the BMI reference of the WHO child growth standards and was updated only for children aged 2–18 years with PEZZ data [8, 9, 20]. Also for head circumference analyses in children aged 0–2 years the two overlapped; both used the head circumference reference from the Zurich Longitudinal Studies [7, 9, 21, 22], and while the CH-WHO 2011 used these references also from 2–18 years, the Swiss 2026 head circumference reference used German data from the KiGGS study from 2–18 years [5]. We used cubic interpolation to obtain LMS values for every age in days from values published in months or years, depending on the reference [1].

To evaluate how well the SwissPedGrowth data fitted to the existing growth references, we calculated mean and standard deviation of z-scores. A perfect fit would yield z-scores following a standard normal distribution (mean 0, SD 1). We interpreted negative or positive deviations of mean z-scores ≥0.50 as strong, 0.25–0.49 as moderate, 0.05–0.24 as weak, and 0–0.04 as negligible [23]. We used penalised cubic B-spline regression to estimate mean z-scores over age.

In the ‘normal growth and weighted’ subsample, we estimated centile curves of height, weight, BMI and head circumference separately for 0–18-year-old boys and girls. We used the LMS method of Cole and Green [24] and estimated Generalized Additive Models for Location Scale and Shape (GAMLSS) using the *gamlss* package in R, following the approach of Stanisopoulos et al. in two steps: first estimating a power transformation of age; second, estimating a model with smooth centile curves [25]. A detailed description can be found in the supplementary material. We drew worm plots (quantile-quantile plots) as diagnostics of the residuals. We selected the model that best balanced model fit (satisfactory residual diagnostics) and centile smoothness. We then compared the SwissPedGrowth centiles visually to those of the Swiss 2026 references.

We used GraphDB Workbench version 10.8.0 to flatten RDF data and R Studio Server version 2024.03.999 with R version 4.3.1 for the statistical analysis. The code for interpolating the growth references and the statistical analysis is available on GitHub (https://github.com/LorenzLeuenberger/Growth-References_SwissPedGrowth).

## RESULTS

### Study population

SwissPedGrowth included 640,170 children with 807,879 height, 2,156,300 weight, 1,341,666 calculated BMI, and 193,091 head circumference recordings at the time of analysis. After excluding children with missing age or sex, and excluding duplicated or biologically implausible anthropometric recordings, the ‘all children’ sample comprised 450,593 (70%) children, 213,868 (33%) with height, 448,002 (70%) with weight, 209,244 (33%) with BMI, and 67,397 (11%) with head circumference recordings (Supplementary Figures S1 to S4). The ‘normal growth and weighted’ subsample comprised 89,315 (14%) children, 39,486 (6%) with height, 77,942 (12%) with weight, 26,971 (4%) with BMI, and 13,007 (2%) with head circumference recordings. Table 1 shows the demographic, socioeconomic, and administrative characteristics of the included samples. The subsample was younger and had more children of Swiss nationality and a lower Swiss-SEP compared with the unweighted ‘all children’ sample. In the subsample, standardised differences for age, sex, nationality, and Swiss-SEP quintiles were small (all <0.1) compared with the general Swiss paediatric population (Supplementary Figure S5).

**Table 1.**
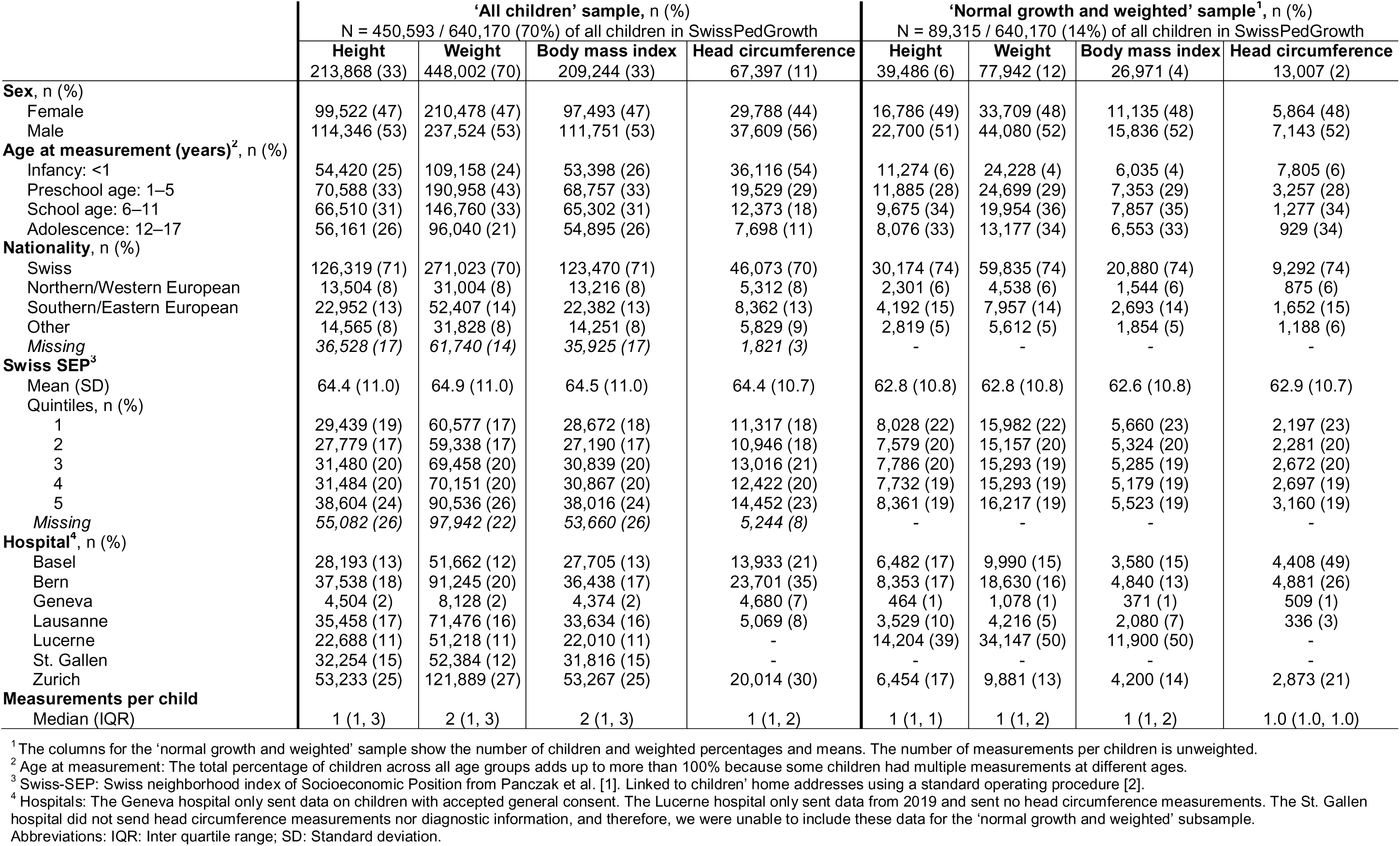
Demographic, socioeconomic, and administrative information of children from the SwissPedGrowth project included in the analysis of height, weight, body mass index, and head circumference.

|  | 'All children' sample, n (%) |  |  |  | 'Normal growth and weighted' sample<sup>1</sup>, n (%) |  |  |  |
| --- | --- | --- | --- | --- | --- | --- | --- | --- |
|  | N = 450,593 / 640,170 (70%) of all children in SwissPedGrowth |  |  |  | N = 89,315 / 640,170 (14%) of all children in SwissPedGrowth |  |  |  |
|  | Height | Weight | Body mass index | Head circumference | Height | Weight | Body mass index | Head circumference |
|  | 213,868 (33) | 448,002 (70) | 209,244 (33) | 67,397 (11) | 39,486 (6) | 77,942 (12) | 26,971 (4) | 13,007 (2) |
| **Sex, n (%)** |  |  |  |  |  |  |  |  |
| Female | 99,522 (47) | 210,478 (47) | 97,493 (47) | 29,788 (44) | 16,786 (49) | 33,709 (48) | 11,135 (48) | 5,864 (48) |
| Male | 114,346 (53) | 237,524 (53) | 111,751 (53) | 37,609 (56) | 22,700 (51) | 44,080 (52) | 15,836 (52) | 7,143 (52) |
| **Age at measurement (years)<sup>2</sup>, n (%)** |  |  |  |  |  |  |  |  |
| Infancy: <1 | 54,420 (25) | 109,158 (24) | 53,398 (26) | 36,116 (54) | 11,274 (6) | 24,228 (4) | 6,035 (4) | 7,805 (6) |
| Preschool age: 1–5 | 70,588 (33) | 190,958 (43) | 68,757 (33) | 19,529 (29) | 11,885 (28) | 24,699 (29) | 7,353 (29) | 3,257 (28) |
| School age: 6–11 | 66,510 (31) | 146,760 (33) | 65,302 (31) | 12,373 (18) | 9,675 (34) | 19,954 (36) | 7,857 (35) | 1,277 (34) |
| Adolescence: 12–17 | 56,161 (26) | 96,040 (21) | 54,895 (26) | 7,698 (11) | 8,076 (33) | 13,177 (34) | 6,553 (33) | 929 (34) |
| **Nationality, n (%)** |  |  |  |  |  |  |  |  |
| Swiss | 126,319 (71) | 271,023 (70) | 123,470 (71) | 46,073 (70) | 30,174 (74) | 59,835 (74) | 20,880 (74) | 9,292 (74) |
| Northern/Western European | 13,504 (8) | 31,004 (8) | 13,216 (8) | 5,312 (8) | 2,301 (6) | 4,538 (6) | 1,544 (6) | 875 (6) |
| Southern/Eastern European | 22,952 (13) | 52,407 (14) | 22,382 (13) | 8,362 (13) | 4,192 (15) | 7,957 (14) | 2,693 (14) | 1,652 (15) |
| Other | 14,565 (8) | 31,828 (8) | 14,251 (8) | 5,829 (9) | 2,819 (5) | 5,612 (5) | 1,854 (5) | 1,188 (6) |
| Missing | 36,528 (17) | 61,740 (14) | 35,925 (17) | 1,821 (3) | - | - | - | - |
| **Swiss SEP<sup>3</sup>** |  |  |  |  |  |  |  |  |
| Mean (SD) | 64.4 (11.0) | 64.9 (11.0) | 64.5 (11.0) | 64.4 (10.7) | 62.8 (10.8) | 62.8 (10.8) | 62.6 (10.8) | 62.9 (10.7) |
| Quintiles, n (%) |  |  |  |  |  |  |  |  |
| 1 | 29,439 (19) | 60,577 (17) | 28,672 (18) | 11,317 (18) | 8,028 (22) | 15,982 (22) | 5,660 (23) | 2,197 (23) |
| 2 | 27,779 (17) | 59,338 (17) | 27,190 (17) | 10,946 (18) | 7,579 (20) | 15,157 (20) | 5,324 (20) | 2,281 (20) |
| 3 | 31,480 (20) | 69,458 (20) | 30,839 (20) | 13,016 (21) | 7,786 (20) | 15,293 (19) | 5,285 (19) | 2,672 (20) |
| 4 | 31,484 (20) | 70,151 (20) | 30,867 (20) | 12,422 (20) | 7,732 (19) | 15,293 (19) | 5,179 (19) | 2,697 (19) |
| 5 | 38,604 (24) | 90,536 (26) | 38,016 (24) | 14,452 (23) | 8,361 (19) | 16,217 (19) | 5,523 (19) | 3,160 (19) |
| Missing | 55,082 (26) | 97,942 (22) | 53,660 (26) | 5,244 (8) | - | - | - | - |
| **Hospital<sup>4</sup>, n (%)** |  |  |  |  |  |  |  |  |
| Basel | 28,193 (13) | 51,662 (12) | 27,705 (13) | 13,933 (21) | 6,482 (17) | 9,990 (15) | 3,580 (15) | 4,408 (49) |
| Bern | 37,538 (18) | 91,245 (20) | 36,438 (17) | 23,701 (35) | 8,353 (17) | 18,630 (16) | 4,840 (13) | 4,881 (26) |
| Geneva | 4,504 (2) | 8,128 (2) | 4,374 (2) | 4,680 (7) | 464 (1) | 1,078 (1) | 371 (1) | 509 (1) |
| Lausanne | 35,458 (17) | 71,476 (16) | 33,634 (16) | 5,069 (8) | 3,529 (10) | 4,216 (5) | 2,080 (7) | 336 (3) |
| Lucerne | 22,688 (11) | 51,218 (11) | 22,010 (11) | - | 14,204 (39) | 34,147 (50) | 11,900 (50) | - |
| St. Gallen | 32,254 (15) | 52,384 (12) | 31,816 (15) | - | - | - | - | - |
| Zurich | 53,233 (25) | 121,889 (27) | 53,267 (25) | 20,014 (30) | 6,454 (17) | 9,881 (13) | 4,200 (14) | 2,873 (21) |
| **Measurements per child** |  |  |  |  |  |  |  |  |
| Median (IQR) | 1 (1, 3) | 2 (1, 3) | 2 (1, 3) | 1 (1, 2) | 1 (1, 1) | 1 (1, 2) | 1 (1, 2) | 1.0 (1.0, 1.0) |

<sup>1</sup> The columns for the 'normal growth and weighted' sample show the number of children and weighted percentages and means. The number of measurements per children is unweighted.
<sup>2</sup> Age at measurement: The total percentage of children across all age groups adds up to more than 100% because some children had multiple measurements at different ages.
<sup>3</sup> Swiss-SEP: Swiss neighborhood index of Socioeconomic Position from Panczak et al. [1]. Linked to children' home addresses using a standard operating procedure [2].
<sup>4</sup> Hospitals: The Geneva hospital only sent data on children with accepted general consent. The Lucerne hospital only sent data from 2019 and sent no head circumference measurements. The St. Gallen hospital did not send head circumference measurements nor diagnostic information, and therefore, we were unable to include these data for the 'normal growth and weighted' subsample.
Abbreviations: IQR: Inter quartile range; SD: Standard deviation.

### Fit of height, weight, BMI, and head circumference to Swiss growth references

The ‘all children’ sample in SwissPedGrowth was slightly shorter (mean height z-score: -0.19), slightly lighter (mean weight z-score: -0.09), had a slightly lower BMI (mean BMI z-score: -0.07), and had a moderately lower head circumference (mean head circumference z-score: -0.28) compared with the Swiss 2026 growth references (Table 2). In the ‘normal growth and weighted’ subsample, mean height (0.00), weight (0.01), BMI (0.05), and head circumference (0.01) z-scores did not deviate from the Swiss 2026 reference. Variability was higher in the ‘all children’ sample for height, weight, BMI, and head circumference compared with both the Swiss 2026 and CH-WHO 2011 references (Figure 1). Although the variability was reduced in the subsample, it was still higher compared with the growth references (SD >1, Table 2).

**Figure 1.**
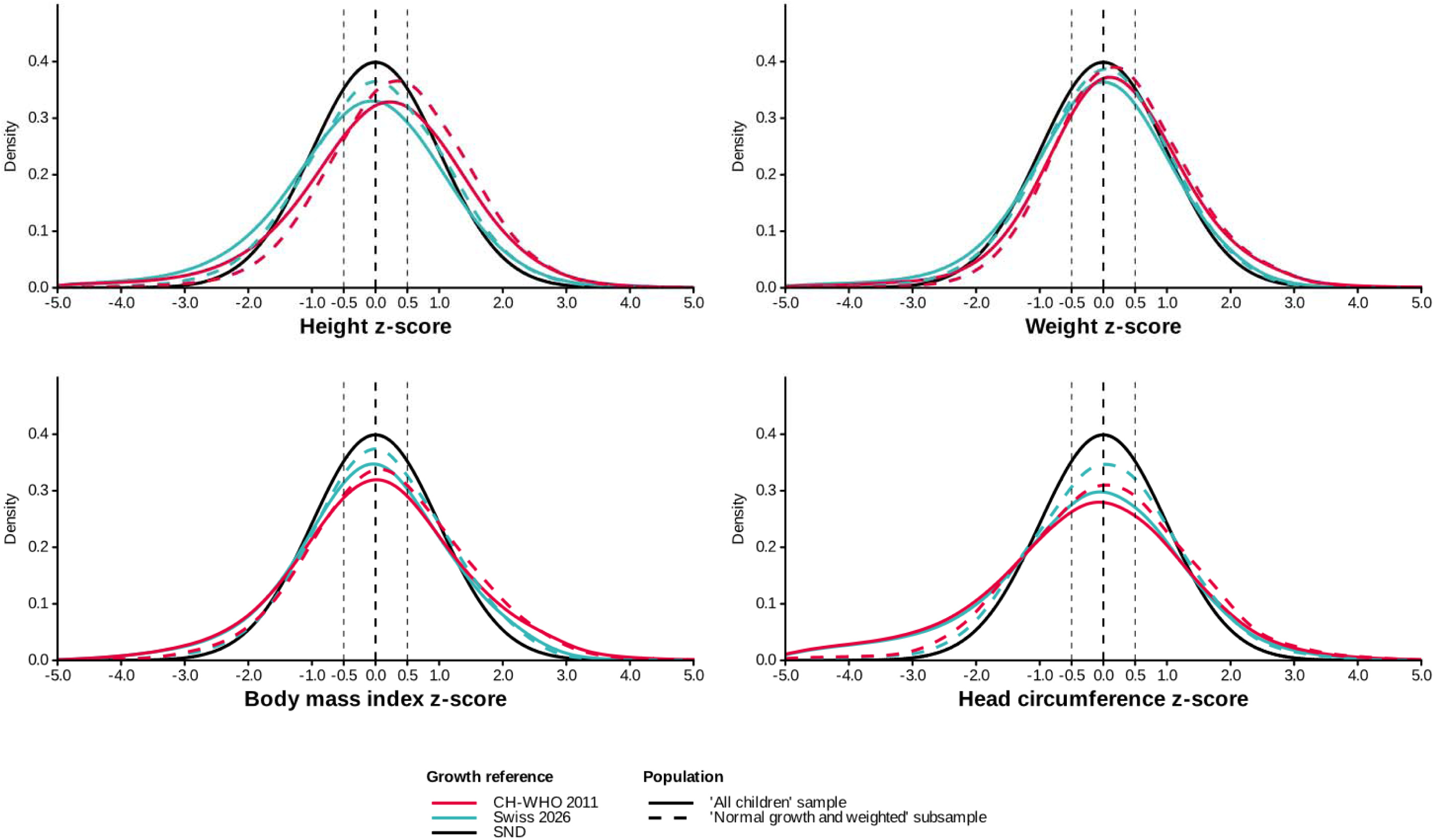
Z-score distributions of height, weight, BMI, and head circumference for children in the SwissPedGrowth project. Z-score distributions based on Swiss 2026 growth references for height, weight, and body mass index and the CH-WHO 2011 growth reference for head circumference are shown versus a standard normal distribution (perfect fit). Abbreviations: CH-WHO 2011: WHO growth references adopted for Switzerland [3]; Swiss 2026: Growth references recommended in 2026 [4, 5]; SND: Standard normal distribution.

**Table 2.** Mean and standard deviation of height, weight, body mass index and head circumference z-scores of participants in the SwissPedGrowth project across growth references used in Switzerland.

|  | Growth references |  |
| --- | --- | --- |
|  | CH-WHO 2011 | Swiss 2026 |
| **Height z-score, mean (SD)** |  |  |
| 'All children' sample | 0.10 (1.32) | -0.19 (1.31) |
| 'Normal growth and weighted' subsample | 0.34 (1.13) | 0.00 (1.13) |
| **Weight z-score, mean (SD)** |  |  |
| 'All children' sample | 0.16 (1.20) | -0.09 (1.21) |
| 'Normal growth and weighted' subsample | 0.27 (1.08) | 0.01 (1.07) |
| **Body mass index z-score, mean (SD)** |  |  |
| 'All children' sample | 0.04 (1.36) | -0.07 (1.24) |
| 'Normal growth and weighted' subsample | 0.18 (1.23) | 0.05 (1.10)<sup>1</sup> |
| **Head circumference z-score, mean (SD)** |  |  |
| 'All children' sample | -0.28 (1.55) | -0.28 (1.49) |
| 'Normal growth and weighted' subsample | 0.04 (1.33) | 0.01 (1.17) |

95% confidence intervals for mean z-scores were $\pm 0.01$ to 0.02 in the 'all children' sample and $\pm 0.01$ to 0.05 in the 'normal growth and weighted' subsample.
<sup>1</sup> The mean BMI z-score in the 'normal growth and weighted' subsample rounded to four decimal digits was 0.0475, below 0.05, wherefore we refer to it in the text as "BMI z-scores did not deviate from the Swiss 2026 references".
Abbreviations: CH-WHO 2011: WHO growth references adopted for Switzerland [3]; Swiss 2026: Growth references recommended in 2026 [4, 5]; SD: Standard deviation.

Mean height z-scores of the ‘all children’ sample based on the Swiss 2026 reference seemed close to zero from ages 1–15 years, with a decrease in both boys and girls aged <1 year and in boys aged >15 years (Figure 2). Mean weight z-scores were also close to zero, with a similar decrease in boys and girls aged <1 year and boys aged >15 years. Mean BMI z-scores were close to zero, with a similar decrease in boys and girls aged <1 year. Mean head circumference z-scores were below zero in boys and girls aged <2 years and again below zero in boys aged 12–18 years. In the subsample, the mean height z-scores were close to zero in children aged <1 year and boys aged >15 years. The mean weight and BMI z-scores still remained below zero in children aged <1 year, similar to the pattern in the ‘all children’ sample.

**Figure 2.**
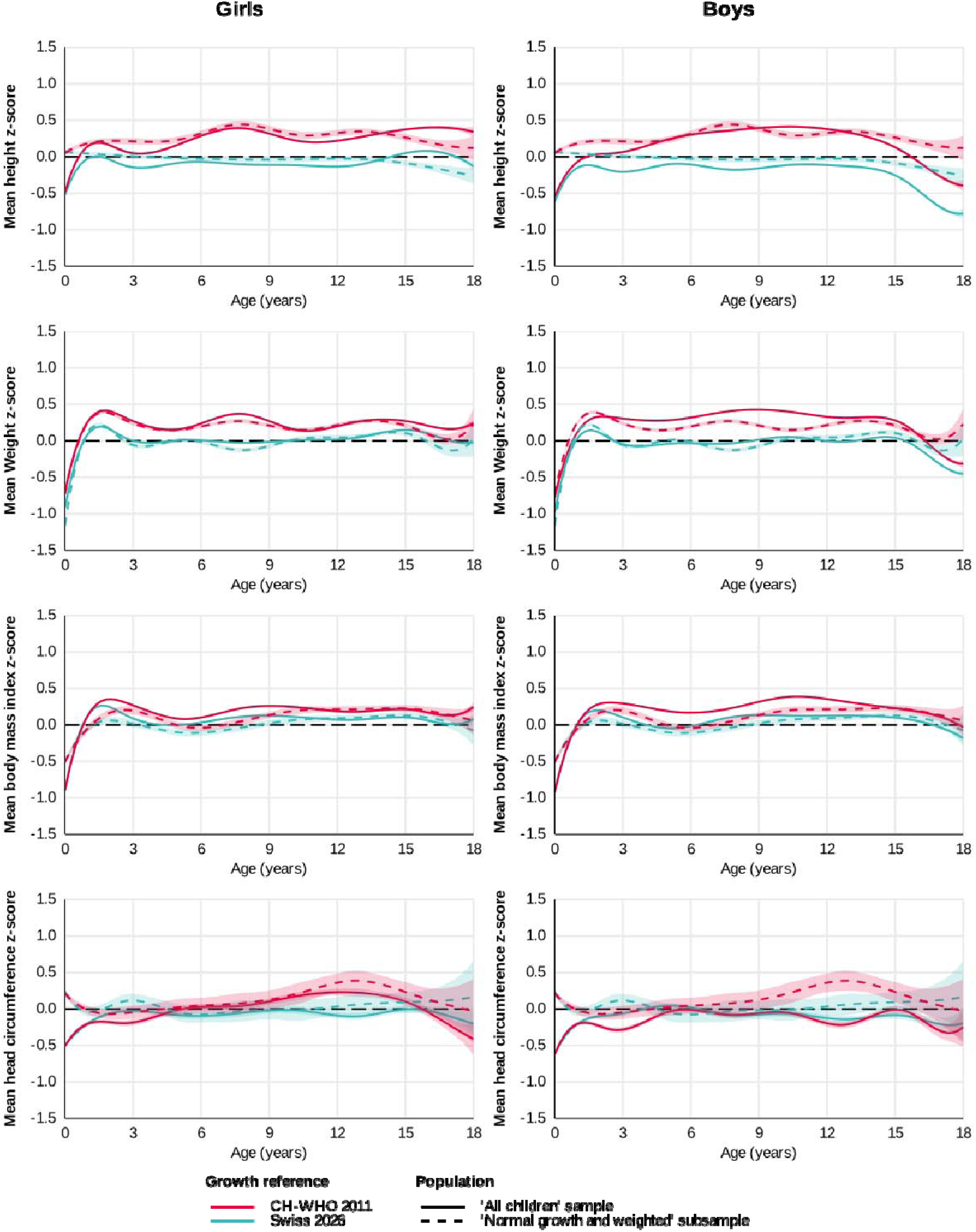
Mean height, weight, body mass index, and head circumference z-scores over age for girls and boys in the SwissPedGrowth project. Mean z-scores based on Swiss 2026 growth references for height, weight, and body mass index and the CH-WHO 2011 growth reference for head circumference were locally estimated by penalised cubic B-spline regression on age. Abbreviations: CH-WHO 2011: WHO growth references adopted for Switzerland [3]; Swiss 2026: Growth references recommended in 2026 [4, 5].

### Estimated centile curves for height, weight, BMI, and head circumference in girls and boys in SwissPedGrowth

For girls, the centile curves estimated from height, weight, and BMI in the ‘normal growth and weighted’ subsample in SwissPedGrowth aligned well with those of the Swiss 2026 references, although with slightly wider spread (Figure 3). The 50^th^ centile of the height curves closely followed that of the Swiss 2026 reference in girls aged 0–18 years. The 3^rd^ and 97^th^ height centiles were approximately 1.5 cm wider than in the Swiss 2026 reference in girls aged 1–4 years, and the 97^th^ centile was up to 4 cm lower in girls aged 14–18 years. The 50^th^ weight centile closely followed that of the Swiss 2026 reference, and was approximately 2 kg higher in girls aged 13–15 years. The 3^rd^ centile was 1 kg lower in girls aged 0–6 months, and the 3^rd^ and 97^th^ centiles were up to 4 kg wider than in the Swiss 2026 reference in girls aged 16–18 years. The 50^th^ BMI centile showed a less pronounced peak in the first year of life compared with that of the Swiss 2026 reference—when the reference was based on the WHO child growth standards—but closely followed it in girls aged 2–9 years and was up to 0.5 kg/m^2^ higher thereafter. The 3^rd^ centile was approximately 0.5 kg/m^2^ lower in girls aged 2–9 years and 16–18 years; the 97^th^ centile was 0.5 kg/m^2^ higher in girls aged 2–5 years and up to 1 kg/m^2^ higher in girls aged 10–18 years. The 50^th^ head circumference centile of SwissPedGrowth closely followed that of the Swiss 2026 reference in girls aged 0–18 years, but the 3^rd^ and 97^th^ centiles were wider spread, up to 1 cm in girls aged 2 years—when the reference was based on the norms of the Zurich Longitudinal Study—and approximately 0.5 cm from 2–18 years—when the reference was based on the German KiGGS data.

**Figure 3.**
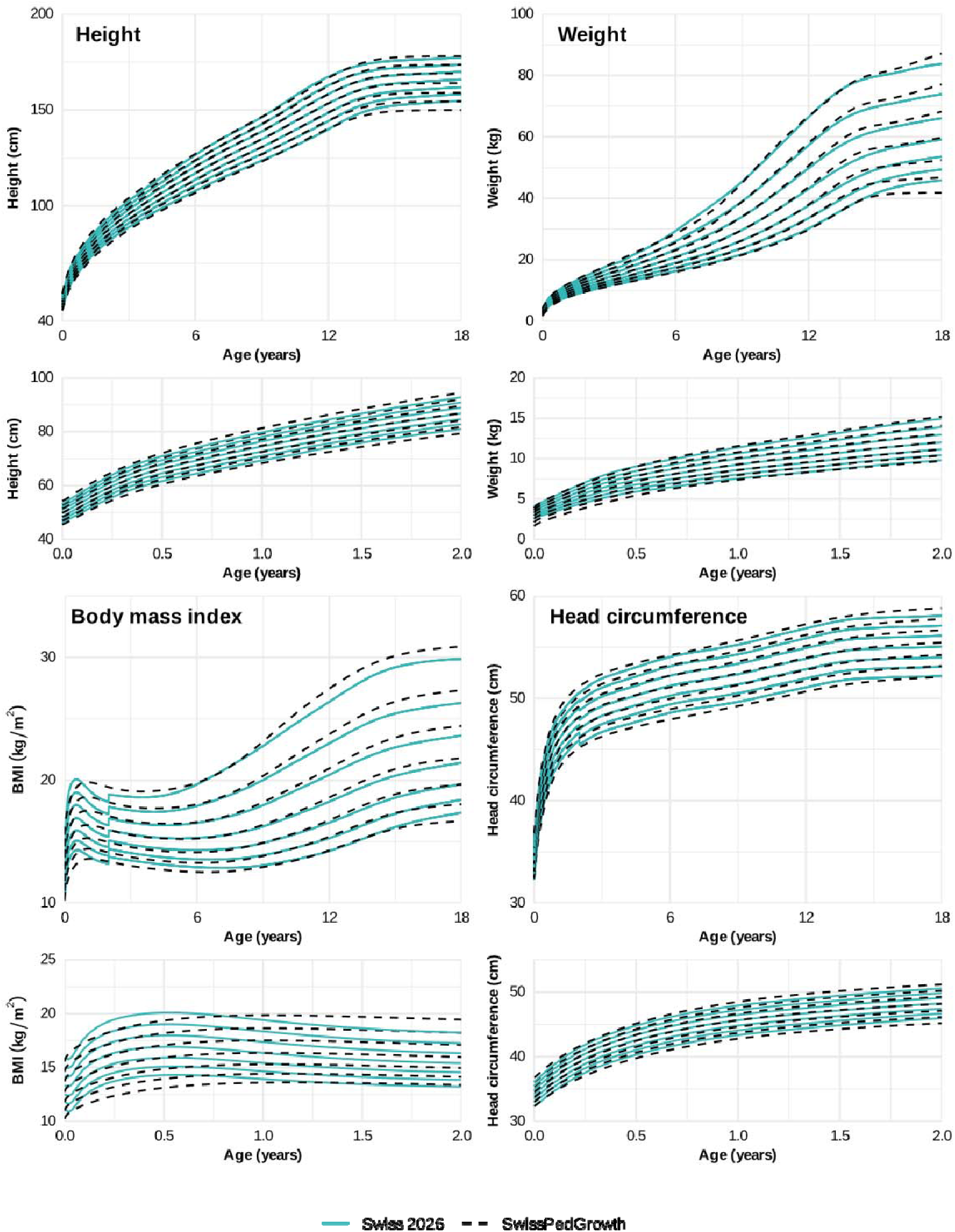
Height, weight, body mass index and head circumference centiles of girls in the SwissPedGrowth project estimated through GAMLSS, ‘normal growth and weighted’ subsample. Shown are the 3^rd^, 10^th^, 25^th^, 50^th^, 75^th^, 90^th^, 97^th^ centiles. The chosen GAMLSS models with good fit (worm plots), parsimony, and smoothness were height: BCCGo, mu restricted to increase monotonically, and nu set to 1, p-splines fitted with GAIC and k = 8; weight: BCCGo, mu restricted to increase monotonically, and a variable nu, p-splines fitted with GAIC and with k = 8; body mass index: BCCGo, mu not restricted to increase monotonically, and a variable nu, p-splines fitted with GAIC and with k = log(n); and head circumference: BCCGo, mu restricted to increase monotonically, and nu set to 1, p-splines fitted with GAIC and with k = 4. The estimated centiles are compared to the Swiss 2026 growth references.

For boys, the centile curves estimated from height, weight, and BMI in the SwissPedGrowth subsample also aligned well with those of the Swiss 2026 references, though again with slightly wider spread (Figure 4). The 50^th^ centile of the height curves closely followed that of the Swiss 2026 reference in boys aged 0–18 years. The 3^rd^ and 97^th^ height centiles were approximately 0.5 to 1 cm wider spread than in the Swiss 2026 reference in boys aged 1–6 years, and the 3^rd^ centile was up to 3 cm lower in boys aged 9–18 years. The 50^th^ weight centile closely followed that of the Swiss 2026, and was approximately 1 kg higher in boys aged 14–16 years. The 3^rd^ centile was 1 kg lower in boys aged 0–6 months and 1 to 4 kg lower in boys aged 10–18 years; the 97^th^ centile was up to 2.5 kg lower in boys aged 16–18 years. The 50^th^ BMI centile showed a less pronounced peak in the first year of life compared with that of the Swiss 2026 reference, but closely followed it in boys aged 2–18 years. The 3^rd^ centile was approximately 1 kg/m^2^ lower in boys aged 16–18 years; the 97^th^ centile was 1 kg/m^2^ higher in boys aged 7–16 years. The head circumference centiles of boys in the subsample aligned well with the CH-WHO 2011 reference, but the 3^rd^ and 97^th^ centiles were wider spread. The 50^th^ head circumference centile closely followed that of the Swiss 2026 reference in boys aged 0–18 years, but the 3^rd^ and 97^th^ centiles were wider spread. The 3^rd^ centile was approximately 1 cm lower in boys aged 2 years and up to 0.5 cm lower from 2–18 years. The 97^th^ centile was maximally 0.5 cm higher.

**Figure 4.**
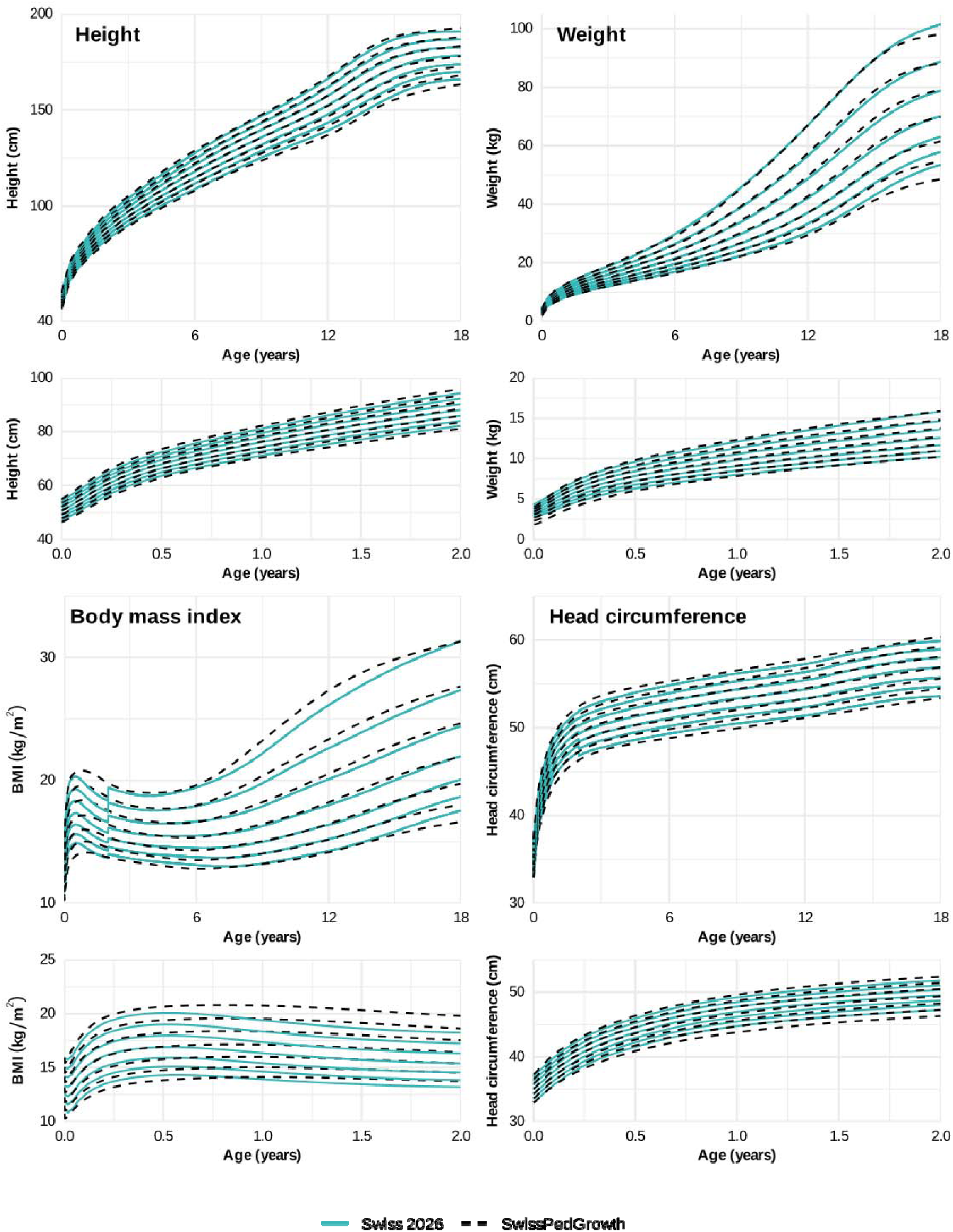
Height, weight, body mass index and head circumference centiles of boys in the SwissPedGrowth project estimated through GAMLSS, ‘normal growth and weighted’ subsample. Shown are the 3^rd^, 10^th^, 25^th^, 50^th^, 75^th^, 90^th^, 97^th^ centiles. The chosen GAMLSS models with good fit (worm plots), parsimony, and smoothness were height: BCCGo, mu restricted to increase monotonically, and nu set to 1, p-splines fitted with default settings; weight: BCCGo, mu restricted to increase monotonically, and a variable nu, p-splines fitted with GAIC and k = 8, we excluded weight measurements at birth (age days = 0) because including those measurements resulted in an unrealistic edge effect due to high variability; body mass index: BCCGo, mu not restricted to increase monotonically, and a variable nu, p-splines fitted with GAIC and k = 6; and head circumference: BCCGo, mu restricted to increase monotonically, and nu set to 1, p-splines fitted with GAIC and with k = 8. The estimated centiles are compared to the Swiss 2026 growth references.

Worm plots showed a good fit of the modelled centiles to the underlying anthropometric data, although they indicated residual skewness in some age groups and a deviation of the median centile of weight in 1–5-year-old girls and <1- and 1–5-year-old boys (Supplementary Figures S6 and S7).

## DISCUSSION

### Principal findings

This study demonstrates that anthropometric data from a large hospital-based paediatric population are highly representative of growth patterns in the general Swiss population. Height, weight, BMI, and head circumference closely aligned with current Swiss growth references. After excluding diseases affecting growth and applying statistical weighting to make the subsample more representative of the general Swiss paediatric population, mean height, weight, and BMI fitted the Swiss 2026 references, and head circumference fitted both the CH-WHO 2011 and Swiss 2026 references. The centile curves estimated from anthropometric data of the subsample of SwissPedGrowth seemed to closely follow the Swiss 2026 references. Whereas the 50^th^ centiles aligned closely, the 3^rd^ and 97^th^ centiles were slightly wider spread, especially in infancy and late adolescence.

### Comparison with existing literature

Previous French studies have demonstrated the feasibility of using EHR data for growth references [2, 26, 27]. Similarly, a recent study from Catalonia, Spain, used routinely collected data to construct growth references for children and adolescents [28]. Both studies extracted EHRs from paediatric primary care practices. To our knowledge, ours is the first study to extract anthropometric EHR data from tertiary care children’s hospitals for evaluating and constructing growth references. Height, weight, BMI, and head circumference of children visiting paediatric hospitals aligned well with the current Swiss 2026 references, which were constructed from primary care data [8], although with somewhat higher variability in the hospital data. Small misalignments with existing centile curves may also result from different modelling approaches. For example, the BMI WHO child growth standards had been modelled separately from 0–2 and from 2–5 years of age and show a distinct peak in the first year of life [20]. In our study, we modelled BMI from 0–18 years and the BMI peak was less pronounced, as described also in the German BMI references [29].

To make hospital EHR data usable for growth research and reduce the higher variability, we carefully cleaned the data, selected a subsample of children without diseases potentially affecting growth, and weighted the study population to better represent the general population. This reduced the increased variability in the hospital data and resulted in a closer fit to the Swiss 2026 references, although variability was still higher and the fit was not perfect in infants and adolescents. This may reflect an overrepresentation of neonates with acute illness or prematurity compared with healthy term-born infants, which is inherent to hospital-based cohorts. Similarly, the higher variability in adolescents aged >15 years may reflect children with chronic diseases still being cared for at paediatric departments, while healthy adolescents may transfer to adult care; usually at age 16 years. We did not extract data from adult hospitals in SwissPedHealth.

In the context of evaluating national and international growth references, a systematic review has shown limited fit of the WHO growth references to local paediatric populations [23]. Accordingly, many Central European countries use local growth references, including Austria [3, 30], France [2, 26, 27], Germany [5], and Italy [4]. Consistent with these findings from neighbouring countries and our previous evaluation in Swiss school children [1], our study shows a better fit of the new local Swiss 2026 references.

### Strengths and limitations

The study has several strengths, including the large sample size, the multicentre design spanning the largest children’s hospitals in French- and German-speaking parts of Switzerland, and the use of structured anthropometric data. SwissPedGrowth includes 640,170 children, representing 26% of the population below 20 years living in Switzerland from 2017–2023 (27 birth cohorts of approximately 90,000 children yield a total of 2,430,000 children). Our evaluation is limited to the French- and German-speaking parts of Switzerland because no children’s hospital from the Italian-speaking part participated in SwissPedGrowth, reducing the national representativeness. However, the impact on the results is likely small, since only 4% of the Swiss paediatric population lives in the Italian-speaking part. We further mitigated this by weighting the study population to represent the age, sex, nationality, and Swiss-SEP distribution of the Swiss paediatric population. Many children lacked height (67%), weight (30%), BMI (67%), or head circumference (89%) recordings, and many also lacked diagnostic information. The ‘normal growth and weighted’ subsample analysis was therefore limited to a smaller sample size (39,486 children with height, 77,942 children with weight, 26,971 children with BMI, and 13,007 children with head circumference recordings). Nevertheless, this remains comparable to the sample size of the Swiss 2026 references (N = 43,290) and consistent with previous simulation studies showing that sample sizes of 7,000–25,000 per sex result in reasonably small standard errors for centiles [8, 31]. In hospitals, anthropometric parameters may be measured less rigorously than in primary care and in research studies on childhood growth, and some recordings in hospitals may even be parent-reported. We mitigated this by using both a self-developed and a pre-existing algorithm to detect duplicates and outliers [11].

### Implications

pädiatrie schweiz (Swiss Society of Paediatrics) announced in 2026 that it recommends the new Swiss 2026 growth references for use in Switzerland. Our evaluation, using more than 540,000 height, 1.4 million weight, 450,000 BMI, and 190,000 head circumference values from Swiss children, suggests that the Swiss 2026 references fitted the hospital data better than the CH-WHO 2011 references, supporting their use in current clinical practice. We also show that anthropometric data from hospitals align well with those from primary care (Swiss 2026 references). This suggests that hospital EHR data are a valid source for not only clinical care and research, but also population health research on childhood growth. Furthermore, EHRs may be a valuable source of anthropometric data for evaluating growth references, or even contributing to their construction, in the future. Particularly, because EHRs may represent a greater social and ethnic heterogeneity than traditional growth studies. However, information about diagnoses and additional demographic and socioeconomic data are necessary to select a population with normal growth and to ensure representativeness of the general population. In settings where adult departments care for adolescents, EHRs from adult hospitals also need to be available to construct growth references up to 18 years. Because prospectively and manually collecting anthropometric data in sufficient quantity to construct reference centiles is logistically demanding with high operational costs (multiple hundred thousand Swiss Francs for study setup and data collection in primary care and school), using EHRs may be particularly attractive for updating growth references more frequently (multiple ten thousand Swiss Francs for reusing EHRs through established frameworks like SwissPedHealth and SwissPedGrowth). Rather than producing static growth references every ten to twenty years, they could be updated continuously and also serve a public health purpose to early identify differences across regions or subpopulations.

### Conclusions

Centile curves estimated from anthropometric data of hospital EHRs aligned well with current Swiss growth references after excluding diseases potentially affecting growth and weighting the study population to the general population. This suggests that hospital EHR data can be used for growth research and may be a suitable source for evaluation, and more frequent updates, of growth references.

## GLOSSARY

EHR: Electronic health records
CDW: Clinical data warehouse
RDF: Resource description framework
SPHN: Swiss Personalized Health Network
SETT: Secure encryption and transfer tool
BMI: Body mass index

## AUTHOR CONTRIBUTIONS

CEK, JAB, RM, FB, and LJS acquired funding. CEK, FB, and LML conceptualised the study. FB and RM set up and coordinated the data extraction. XD, NG, FB, RM, YS, and LML contributed to data curation. LML conducted the formal analysis with support from YS, FR, and MS, under the supervision of FB, BS, and CEK. MAB, CS, VS, FRG, MH, PM, and OGJ validated criteria to exclude anthropometric outliers and diagnoses potentially affecting anthropometric recordings. All authors interpreted and discussed the findings. LML wrote the original draft, and all authors critically reviewed and edited the manuscript. All authors approved the final version of the manuscript.

## DATA AVAILABILITY

Data may be made available to investigators upon request by email to the corresponding author. Metadata of SwissPedHealth can be explored in the metadata catalogue of the Swiss Personalized Health Network: https://schemascope.dcc.sib.swiss/.

## ACKNOWLEDGEMENTS

We thank pädiatrie schweiz for kindly providing the detailed LMS values of the CH-WHO 2011 growth references from Braegger et al. [7]. We thank Tim Aeppli for reviewing the list of diseases potentially affecting anthropometric recordings. We thank Mark Saadeh for manuscript editing.

SwissPedHealth consortium: Andrea Agostini, Anita Rauch, Anna Hartung, Audrey van Drogen, Aurélie Martin Necker, Ben D Spycher, Christian Kahlert, Christopher B Forrest, Claudia E Kuehni, Cornelia Hagman, D Sean Froese, Daphné Chopard, Dylan Lawless, Effy Vayena, Eirini I Petrou, Emanuele Palumbo, Eric Giannoni, Fabiën N Belle, Ioannis Xenarios, Jacques Fellay, Jana Pachlopnik Schmid, Johannes Trück, Julia A Bielicki, Julia E Vogt, Kathrin Hofmann, Katrin Männik, Kelly Ormond, Klara Posfay-Barbe, Lorenz M Leuenberger, Luregn J Schlapbach, Manon Jaboyedoff, Mariam Ait Oumelloul, Martin Stocker, Matthias R Baumgartner, Nicola Zamboni, Nicole Goebel, Patrick G A Pedrioli, Philipp Latzin, Rebeca Mozun, Sandra Goetze, Seraina Prader, Sebastian Kerzel, Simon Boutry, Suliman Bouizaguen, Sven Schulzke, Tatjana Welzel, Tabitha Arn-Roth, Thomas M Sutter, Varvara Dimopoulou, Vito Rt Zanotelli, Xeni Deligianni, Xenia Bovermann, and Yara Shoman.

