## Supplementary-materials for "Towards Electronic Health Records-Based Paediatric Growth References: Results from the SwissPedGrowth Project"

### Supplementary Methods

#### Data extraction and preparation

The data extraction and preparation steps of SwissPedGrowth have been described previously [1]. Hospitals extracted sociodemographic, administrative, anthropometric, and clinical data from EHRs and mapped them to the SPHN Resource Description Framework (RDF) schema [2]. Hospitals also linked the Neighbourhood Index of Socioeconomic Position (Swiss-SEP) to the patient’s home address using a standard operating procedure [3], then removed the address after linkage. The Swiss-SEP is an area-based socioeconomic index derived from information on education, occupation, rent, and overcrowding of households from national censuses conducted from 2012–2015 [4]. Scores range from 0–100, with higher values indicating a better socioeconomic position. Hospitals transferred datasets to BioMedIT, a secure cluster for processing and analysing medical data.

We cleaned the anthropometric data using a self-developed algorithm and the pre-existing growthcleanr package in R [1, 5]: We identified and corrected unit errors, decimal errors, and swapped recordings (height recorded as weight and vice versa), and excluded duplicate recordings (same-day measurements or values carried forward from previous visits), biologically implausible outliers (height and head circumference z-scores <-5 or >5; weight and body mass index z-scores <-5 or >8), and invalid recordings (zero or negative value, negative age). To create a dataset with BMI values, we matched the closest height recording to each weight recording, allowing for a maximum interval of 30 days, as previously described [1, 6]. We calculated BMI as weight[kg]/height[cm]^2^.

We grouped nationality as Swiss, Northern/Western European, Southern/Eastern European, and Other (non-European). We categorised International Classification of Disease 10^th^ version (ICD-10) diagnoses according to their potential influence on anthropometric parameters. Each diagnostic code was evaluated for whether it might affect height, weight, BMI, head circumference, or all parameters, and for how long the corresponding measurements required exclusion: for example, weight and BMI recordings were excluded for 4 weeks following a gastroenteritis diagnosis, whereas all height, weight, BMI, and head circumference recordings were excluded for children with severe chronic diseases such as cystic fibrosis. This classification was based on exclusion criteria from previous growth studies and the expert opinion of a panel of paediatric endocrinologists, paediatric gastroenterologists, and paediatric developmental specialists (Supplementary Table S1) [7, 8].

#### Modelling centile curves using GAMLSS

In the ‘normal growth and weighted’ subsample, we estimated centile curves of height, weight, BMI and head circumference separately for 0–18-year-old boys and girls. We used the LMS method of Cole and Green [9] and estimated Generalized Additive Models for Location Scale and Shape (GAMLSS) using the *gamlss* package in R, following the approach of Stanisopoulos et al. in two steps: first estimating a power transformation of age; second, estimating a model with smooth centile curves [10]. In the first step, we used the lms function of the *gamlss* package to estimate the power of the age transformation. In the second step, we used the gamlss function to fit models with default settings and, additionally, by minimising the Generalised Akaike information criterion (GAIC) for given distributions and a given penalty k. We specified models with the following distributions: Box-Cox Cole and Green original (BCCGo), with nu = 1 (no skewness) or a variable nu, and either Box-Cox Power Exponential original (BCPEo) or Box-Cox t original (BCTo) based on the output of the lms function. We fitted models iteratively, first with default settings and then by increasing the penalty k in increments of 2, from k = 2 (AIC) to k = log n (BIC). We drew worm plots (quantile-quantile plots) as diagnostics of the residuals. We selected the model that best balanced model fit (satisfactory residual diagnostics) and centile smoothness; where models based on different distributions yielded comparably good fit and smooth centiles, we selected the simpler distribution, e.g., BCCGo nu = 1 rather than BCCGo with a variable nu parameter. We then compared the SwissPedGrowth centiles visually to those of the Swiss 2026 references.

### Supplementary Figures

**Excluded height values**

n = 10,746 / 807,879 (1%), ≥18 years
n = 218,145 / 807,879 (27%), duplicate

n = 31,108 / 807,879 (4%), outlier

n = 166 / 807,879 (<1%), invalid information

n = 2 / 807,879 (<1%), missing sex

**Excluded children**

n = 426,301 / 640,170 (67%), no height recording

n = 1 / 640,170 (<1%), missing sex

**‘All children’ sample**

N = 213,868 / 640,170 (33%)

**Height values**

n = 547,712 / 807,879 (68%)

**Children in SwissPedGrowth**

N = 640,170

**Height values**
n = 807,879

**Excluded height values**

n = 152,774 / 807,879 (19%), disease affecting height

n = 333,754 / 807,879 (41%), no diagnostic information

n = 1,421 / 807,879 (<1%), missing nationality

n = 6,429 / 807,879 (<1%), missing Swiss-SEP

**Excluded children**

n = 168,532 / 640,170 (26%), no height after excluded diagnoses

n = 1,056 / 640,170 (<1%), missing nationality

n = 4,794 / 640,170 (<1%), missing Swiss-SEP

**‘Normal growth and weighted’ subsample**

N = 39,486 / 640,170 (6%)

**Height values**

n = 53,334 / 807,879 (7%)

**Supplementary Figure S1: Flow diagram of children and height values extracted from electronic health records of SwissPedGrowth hospitals included in this analysis.**
We excluded anthropometric recordings from children ≥18 years, recordings with invalid information (negative age, zero or negative value), duplicates (same day or carried forward from previous visit), and biologically implausible outliers flagged by a self-developed algorithm (height and head circumference z-score <-5 or >5; weight and BMI z-score <-5 or >8) or the existing growthcleanr algorithm [5]. We excluded children with missing sex or a missing anthropometric value. For the ‘normal growth and weighted’ subsample, we further excluded height recordings that might have been affected by a disease. Some children had no height recording remaining after these exclusions and were therefore removed from the sample. Abbreviations: Swiss-SEP: Swiss Neighbourhood Index of Socioeconomic Position [4].

**Excluded weight values**

n = 15,841 / 2,156,300 (<1%), ≥18 years

n = 654,080 / 2,156,300 (30%), duplicate

n = 56,342 / 2,156,300 (3%), outlier
n = 202 / 2,156,300 (<1%), invalid information

n = 7 / 2,156,300 (<1%), missing sex

**Excluded children**

n = 192,164 / 640,170 (30%), no weight recording

n = 4 / 640,170 (<1%), missing sex

**‘All children’ sample**

N = 448,002 / 640,170 (70%)

**Weight values**

n = 1,429,828 / 2,156,300 (66%)

**Children in SwissPedGrowth**

N = 640,170

**Weight values**
n = 2,156,300

**Excluded weight values**

n = 402,953 / 2,156,300 (19%), disease affecting weight

n = 857,386 / 2,156,300 (40%), missing diagnostic information

n = 5,854 / 2,156,300 (<1%), missing nationality

n = 15,816 / 2,156,300 (<1%), missing Swiss-SEP

**Excluded children**

n = 360,536 / 640,170 (56%), no weight after excluded diagnoses

n = 2,400 / 640,170 (<1%), missing nationality

n =7,124 / 640,170 (1%), missing Swiss-SEP

**‘Normal growth and weighted’ subsample**

N = 77,942 / 640,170 (12%)

**Weight values**

n = 147,819 / 2,156,300 (7%)

**Supplementary Figure S2: Flow diagram of children and weight values extracted from electronic health records of SwissPedGrowth hospitals included in this analysis.**
We excluded anthropometric recordings from children ≥18 years, recordings with invalid information (negative age, zero or negative value), duplicates (same day or carried forward from previous visit), and biologically implausible outliers flagged by a self-developed algorithm (height and head circumference z-score <-5 or >5; weight and BMI z-score <-5 or >8) or the existing growthcleanr algorithm [5]. We excluded children with missing sex or a missing anthropometric value. For the ‘normal growth and weighted’ subsample, we further excluded weight recordings that might have been affected by a disease. Some children had no weight recording remaining after these exclusions and were therefore removed from the sample. Abbreviations: Swiss-SEP: Swiss Neighbourhood Index of Socioeconomic Position [4].

**Excluded body mass index values**

n = 13,179 / 1,341,666 (<1%), ≥18 years

n = 477,827 / 1,341,666 (36%), duplicate

n = 73,557 / 1,341,666 (5%), outlier
n = 164 / 1,341,666 (<1%), invalid information
n = 3 / 1,341,666 (<1%), missing sex

**Excluded children**

n = 430,925 / 640,170 (67%), no body mass index recording

n = 1 / 640,170 (<1%), missing sex

**‘All children’ sample**

N = 209,244 / 640,170 (33%)

**Body mass index values**

n = 776,936 / 1,341,666 (58%)

**Children in SwissPedGrowth**

N = 640,170

**Body mass index values**
n = 1,341,666

**Excluded body mass index values**

n = 352,524 / 1,341,666 (26%), disease affecting body mass index

n = 370,132 / 1,341,666 (28%), missing diagnostic information

n = 1,237 / 1,341,666 (<1%), missing nationality

n = 5,679 / 1,341,666 (<1%), missing Swiss-SEP

**Excluded children**

n = 178,487 / 640,170 (28%), no body mass index after excluded diagnoses

n = 694 / 640,170 (<1%), missing nationality

n = 3,092 / 640,170 (<1%), missing Swiss-SEP

**‘Normal growth and weighted’ subsample**

N = 26,971 / 640,170 (4%)

**Body mass index values**

n = 47,364 / 1,341,666 (4%)

**Supplementary Figure S3: Flow diagram of children and BMI values extracted from electronic health records of SwissPedGrowth hospitals included in this analysis.**
We excluded anthropometric recordings from children ≥18 years, recordings with invalid information (negative age, zero or negative value), duplicates (same day or carried forward from previous visit), and biologically implausible outliers flagged by a self-developed algorithm (height and head circumference z-score <-5 or >5; weight and BMI z-score <-5 or >8) or the existing growthcleanr algorithm [5]. We excluded children with missing sex or a missing anthropometric value. For the ‘normal growth and weighted’ subsample, we further excluded BMI recordings that might have been affected by a disease. Some children had no BMI recording remaining after these exclusions and were therefore removed from the sample. Abbreviations: BMI: Body mass index; Swiss-SEP: Swiss Neighbourhood Index of Socioeconomic Position [4].

**Excluded head circumference values**

n = 435 / 193,091 (<1%), ≥18 years

n = 34,625 / 193,091 (18%), duplicate

n = 14,268 / 193,091 (7%), outlier
n = 167 / 193,091 (<1%), invalid information

**Excluded children**

n = 572,773 (89%), no head circumference recording

**‘All children’ sample**

N = 67,397 / 640,170 (11%)

**Head circumference values**

n = 143,596 / 193,091 (74%)

**Children in SwissPedGrowth**

N = 640,170

**Head circumference values**
n = 193,091

**Excluded head circumference values**

n = 66,701 / 193,091 (35%), disease affecting head circumference

n = 59,415 / 193,091 (31%), missing diagnostic information

n = 378 / 193,091 (<1%), missing nationality

n = 1,120 / 193,091 (<1%), missing Swiss-SEP

**Excluded children**

n = 53,124 / 640,170 (8%), no head circumference after excluded diagnoses

n = 305 / 640,170 (<1%), missing nationality

n = 961 / 640,170 (<1%), missing Swiss-SEP

**‘Normal growth and weighted’ subsample**

N = 13,007 / 640,170 (2%)

**Head circumference values**

n = 15,982 / 193,091 (8%)

**Supplementary Figure S4: Flow diagram of children and head circumference values extracted from electronic health records of SwissPedGrowth hospitals included in this analysis.**
We excluded anthropometric recordings from children ≥18 years, recordings with invalid information (negative age, zero or negative value), duplicates (same day or carried forward from previous visit), and biologically implausible outliers flagged by a self-developed algorithm (height and head circumference z-score <-5 or >5; weight and BMI z-score <-5 or >8) or the existing growthcleanr algorithm [5]. We excluded children with missing sex or a missing anthropometric value. For the ‘normal growth and weighted’ subsample, we further excluded head circumference recordings that might have been affected by a disease. Some children had no head circumference recording after these exclusions and were therefore removed from the sample. Abbreviations: Swiss-SEP: Swiss Neighbourhood Index of Socioeconomic Position [4].

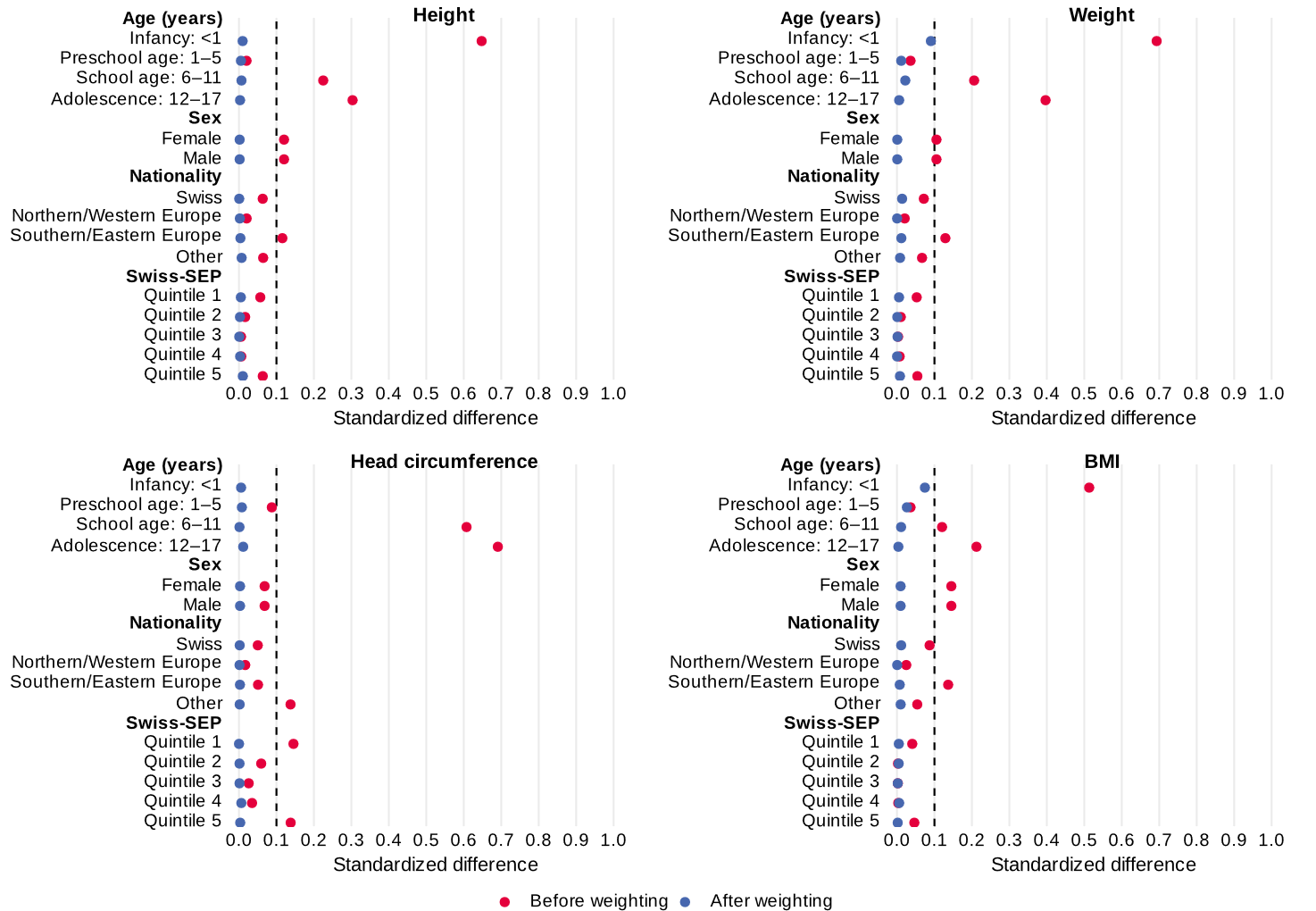

**Supplementary Figure S5. Standardised differences in age, sex, nationality, and Swiss-SEP between the SwissPedGrowth ‘normal growth and weighted’ subsample (height, weight, BMI, and head circumference analyses) and the general paediatric population of Switzerland before and after weighting.**
We used iterative proportional fitting (raking) to weight each subsample to match the age, sex, nationality, and Swiss-SEP distributions of the general population. Standardised differences were calculated using Cohen’s h [11]. Abbreviations: Swiss-SEP: Swiss Neighbourhood Index of Socioeconomic Position [4].

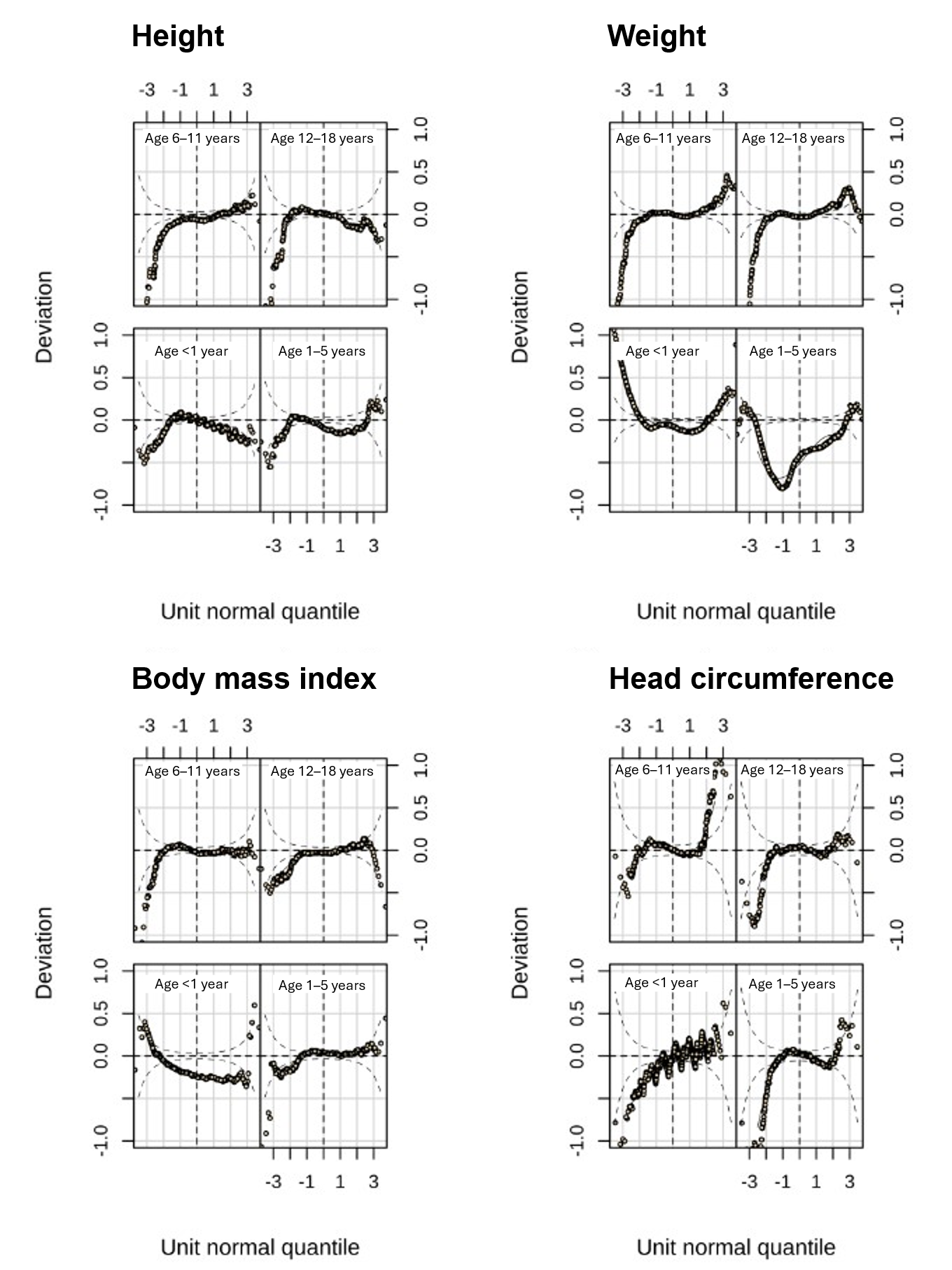

**Supplementary Figure S6. Worm plots for height, weight, body mass index, and head circumference centiles of girls in the SwissPedGrowth ‘normal growth and weighted’ subsample.**
We constructed centiles from the SwissPedGrowth project using the LMS method of Cole and Green and the R package *gamlss* [9, 12]. Each worm plot shows the unit normal quintiles for four age groups: bottom left (<1 year), bottom right (1–5 years), top left (6–11 years), top right (12–18 years).

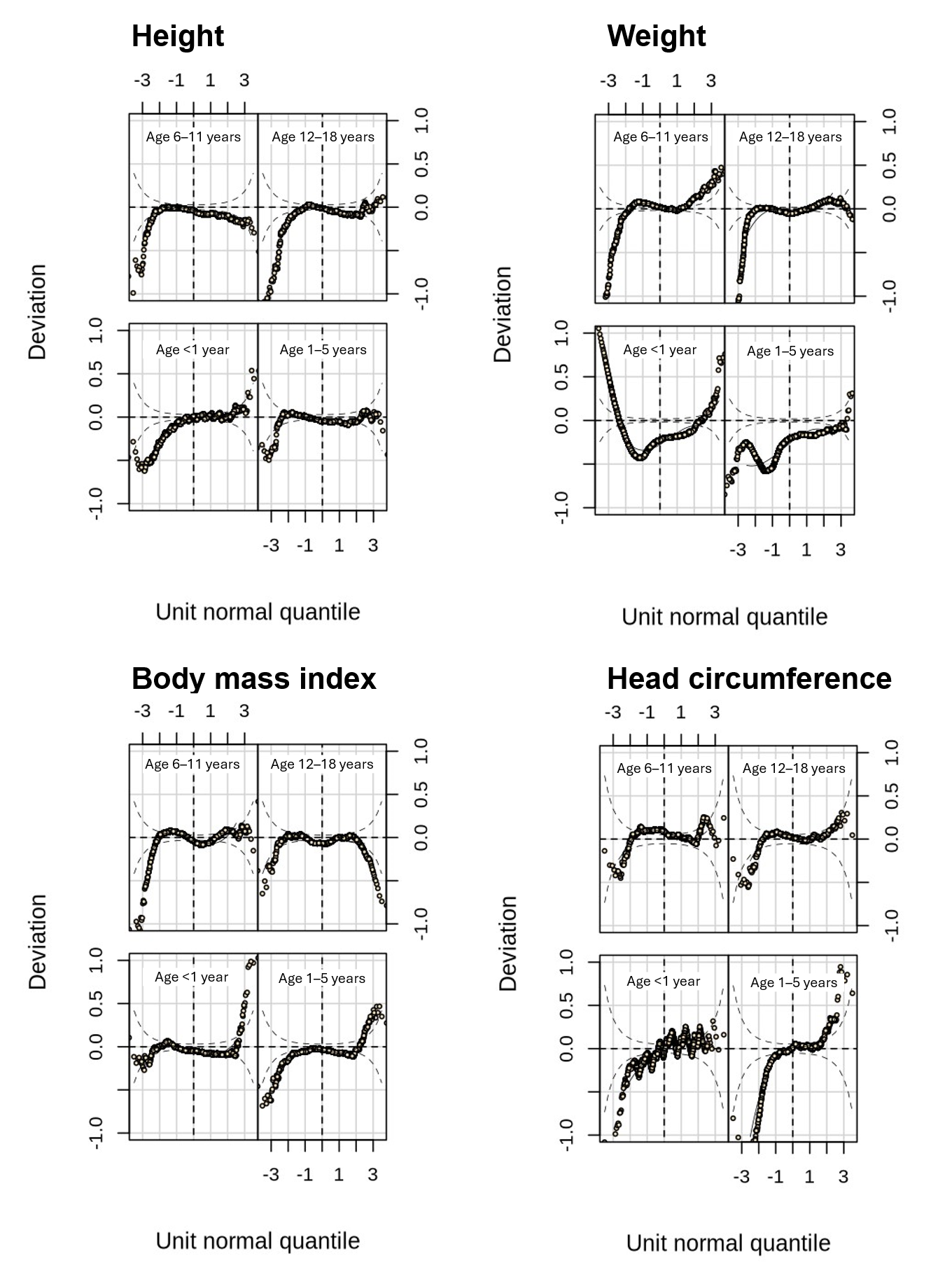

**Supplementary Figure S7. Worm plots for height, weight, body mass index and head circumference centiles of boys in the SwissPedGrowth ‘normal growth and weighted’ subsample.**
We constructed centiles from the SwissPedGrowth project using the LMS method of Cole and Green and the R package *gamlss* [9, 12]. Each worm plot shows the unit normal quintiles for four age groups: bottom left (<1 year), bottom right (1–5 years), top left (6–11 years), top right (12–18 years).
